# Controlling multiple COVID-19 epidemic waves: an insight from a multi-scale model linking the behavior change dynamics to the disease transmission dynamics

**DOI:** 10.1101/2021.04.07.21255056

**Authors:** Biao Tang, Weike Zhou, Xia Wang, Hulin Wu, Yanni Xiao, Sanyi Tang

## Abstract

COVID-19 epidemics exhibited multiple waves regionally and globally since 2020. It is important to understand the insight and underlying mechanisms of the multiple waves of COVID-19 epidemics in order to design more efficient non-pharmaceutical interventions (NPIs) and vaccination strategies to prevent future waves. We propose a multi-scale model by linking the behavior change dynamics to the disease transmission dynamics to investigate the effect of behavior dynamics on COVID-19 epidemics using the game theory. The proposed multi-scale model was calibrated and key parameters related to disease transmission dynamics and behavioral dynamics with/without vaccination were estimated based on COVID-19 epidemic data and vaccination data. Our modeling results demonstrate that the feedback loop between behavior changes and COVID-19 transmission dynamics plays an essential role in inducing multiple epidemic waves. We find that the long period of high-prevalence or persistent deterioration of COVID-19 epidemics could drive almost all population to change their behaviors and maintain the altered behaviors, however, the effect of behavior changes faded out gradually along the progress of epidemics. This suggests that it is essential not only to have persistent, but also effective behavior changes in order to avoid subsequent epidemic waves. In addition, our model also suggests the importance to maintain the effective altered behaviors during the initial stage of vaccination, and to counteract relaxation of NPIs, it requires quick and massive vaccination to avoid future epidemic waves.

## Introduction

The COVID-19 pandemic, a massive global health crisis, has been bringing a great threat to the global health. Till March 8, 2021, the new coronavirus (SARS-CoV-2) has spread to over 200 countries with 116,135,492 confirmed cases and 2,581,976 deaths [1]. Since December 2019, many regions and countries have experienced multiple epidemic waves or new outbreaks even with strong non-pharmaceutical interventions (NPIs) such as lockdown and keeping social distance [1]. It is suspected that the multiple epidemic waves were presumably due to intermittently lifting the lockdown in order to revive the economy. Therefore, the NPIs have been re-enhanced to avoid new waves by increasing hand washing, reducing face touching, wearing masks in public and physical distancing [2-4], which have been demonstrated to be effective in mitigating the new waves of COVID-19 pandemic [5-8].

It is known that this public health crisis has required large-scale behavior changes and placed significant psychological burdens on individuals [9-12]. As pointed out by Ferguson [13], the prevalence or perceived risk-driven behavior change plays an important role in the spread of infectious diseases. In particular, the study by Manfredi and d’Onofrio [14] showed that human behavior change might be a critical explaining factor for oscillations in infectious disease endemics. A number of models have been formulated to investigate the interaction between behavior changes and resurgence of infections [15-19]. Game theory was widely used to formulate the dynamics of human behavioral changes, by assuming that individuals would make the best decisions based on a trade-off between two different strategies, adopting normal or altered behaviors [18-20].

Recently, there are several studies that examined the impact of behavioral changes on COVID-19 epidemics, by assuming a constant proportion of the population choosing to change their behaviors [21, 22], or including the reduced contact rate and/or increased the quarantine rate [23]. In this study, we propose a new multi-scale model to quantify the co-evolution of behavior changes and COVID-19 epidemics to reveal the insight and underlying mechanisms of multiple epidemic waves. Using the concept of game theory for cost-benefit consideration of alternative decisions, we further identified the key factors of behavior changes to mitigate the epidemics, and finally we provide the quantitative guidance for relieving NPIs with continuous vaccination based the established model.

## Main results

### 2.1 Co-evolution model and model fitting

We propose a co-evolution model linking the disease transmission dynamics to the behavioral change dynamics (SI Fig.2) [18-20]. Transmission dynamics follows a compartmental scheme (SEIAHR model for COVID-19), where individuals are divided into susceptible individuals (*S*), exposed individuals *(E)*, infectives with symptoms *(I)*, asymptomatic infectives (*A*), hospitalized individuals *(H*) and recovered individuals (symptomatic recovered individuals, R_I_, and asymptomatic individuals, *R*_*A*_). We model the dynamics of behavioral change, which is triggered by the scare of infection of susceptible individuals, asymptomatic infected individuals and recovered individuals who did not experience symptoms. Let *M(t)* be an information variable (or perceived infection) governing the signal available to individuals as a function of infections (or particular confirmed cases), *B(t)* represent the fractions of individuals performing no behavior changes in term of perceived infections [18,19].

Note that in our modelling framework, Model (1), there are four key factors related to behavioral change dynamics: *m*, the sensitivity of individuals to perceived infection (a higher value of *m* indicating the higher sensitivity); η, the speed of raising the risk awareness; v, the persistence to maintain the risk awareness; ρ, the spread rate of the behavior changes among individuals. Further, as the behavior changed, q denotes the reduced transmission rate due to behavioral changes. Intervention measures were modeled through modifications of the diagnosis rate (continuous function), disease-induced death rate (piece-wise function of time), behavioral change-related parameters (*q, η, ρ*) as piece-wise functions of time accounting for the variation in interventions at different epidemic phases.

The least squares (LS) method was used to fit the proposed model and the bootstrap approach was used to obtain the 95% confidence interval (Fig. 1), and the estimated parameters and their standard deviations are listed in Table 1. In particular, the empirical distributions of the estimated key parameters for behavioral changes, *(q, ρ, η,v,m)*, for Hongkong and the world from the bootstrap method (500 bootstrap samples), are shown in SI Fig. 3. We also obtained the estimate of the effective reproduction numbers and their confidence intervals for the three regions/countries (Hongkong, Japan, USA) and the world based on the bootstrap method (Fig. 1). The multi-scale model is proposed to capture the transmission dynamics of COVID-19 epidemics as well as the evolution of behavioral changes, and the model was calibrated based on COVID-19 epidemic data from multiple sources, and then used to evaluate the impact of enhancing/relieving NPIs upon vaccination. More details are available in the section of Methods.

**Fig 1.**
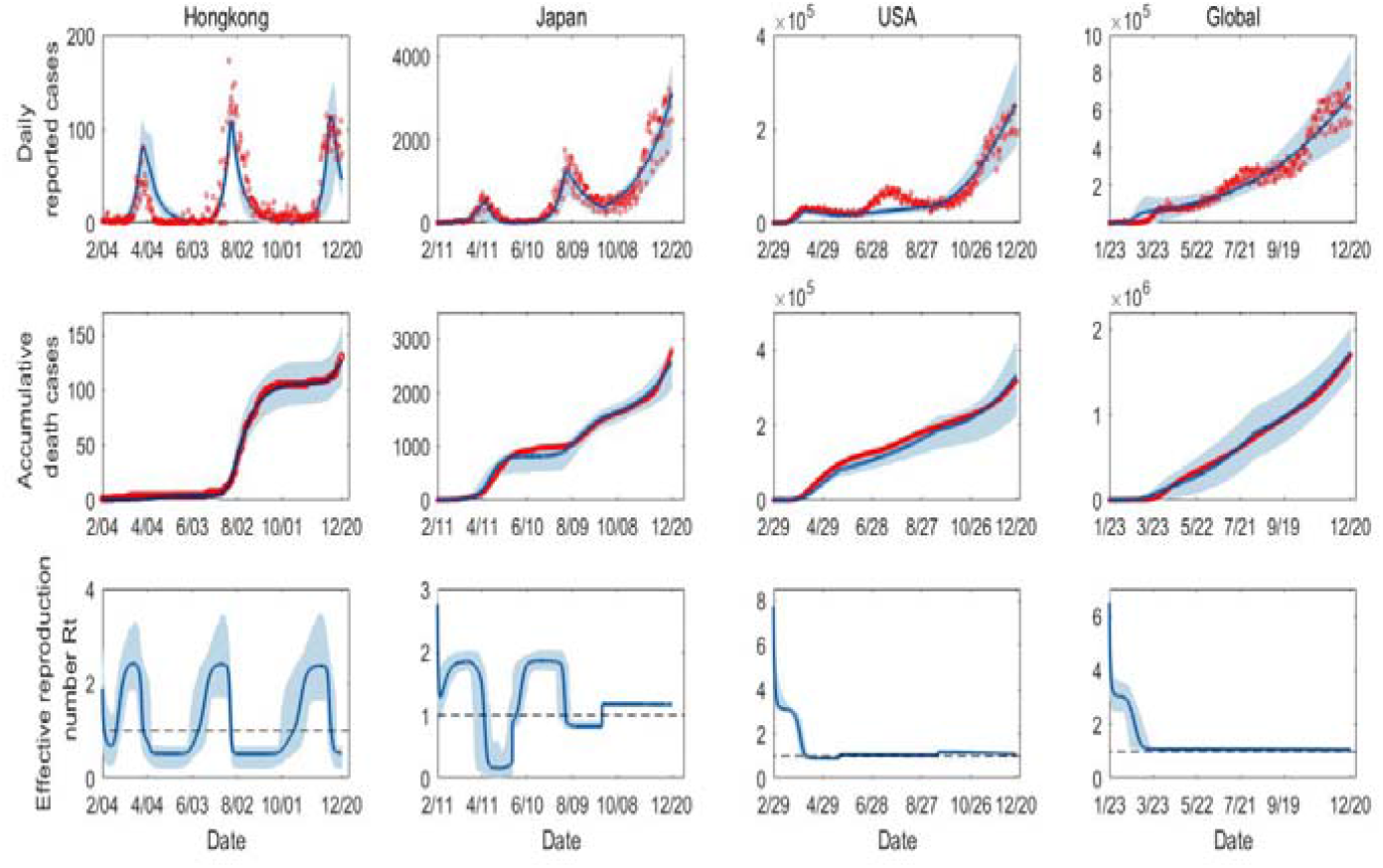
Model fitting results for Model (1): The blue curves are the estimated curves with the shadow areas as the corresponding 95% confidence band. The red cycles are the observed data of the daily reported cases and the accumulative death cases

**Table 1.**
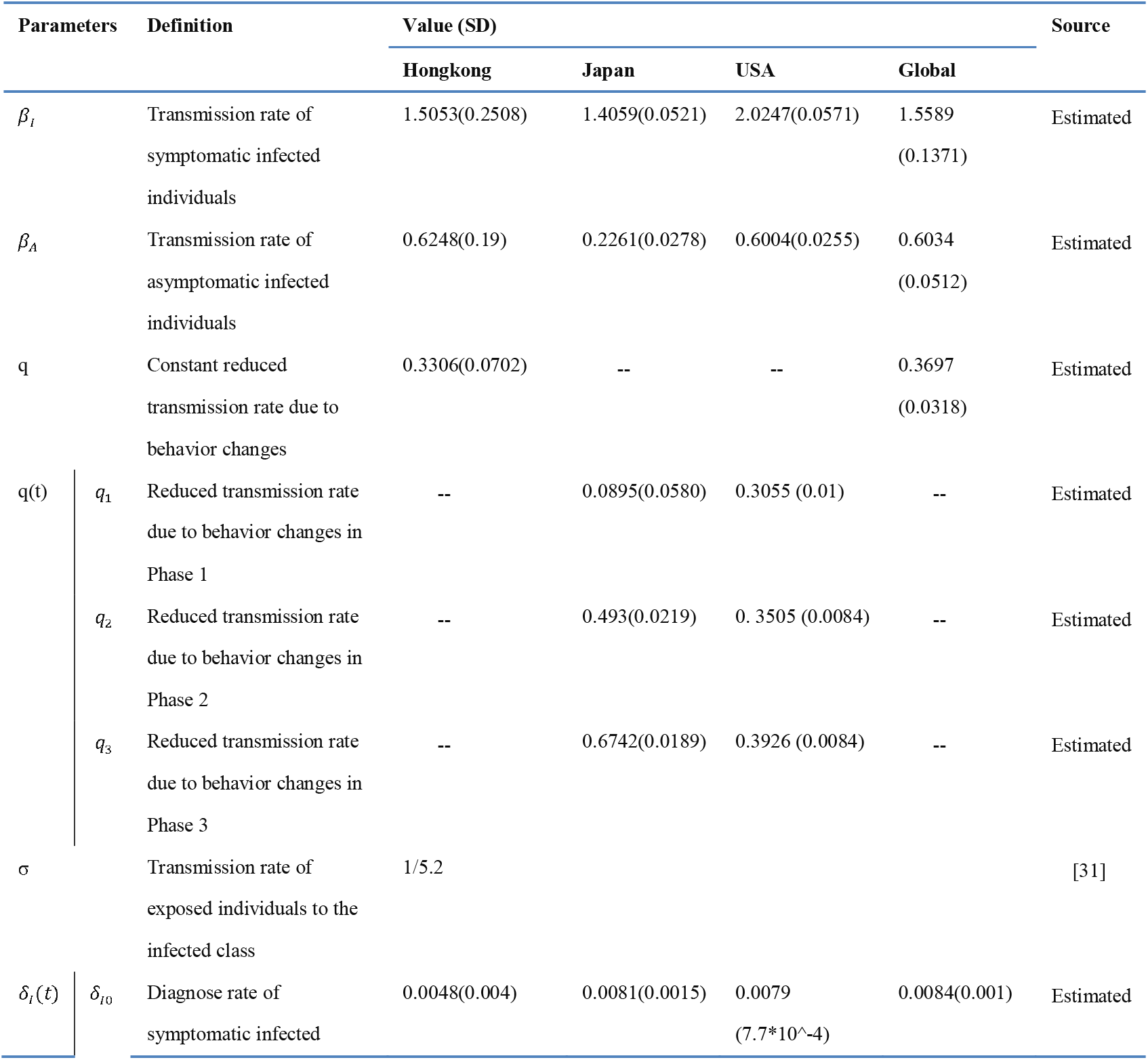
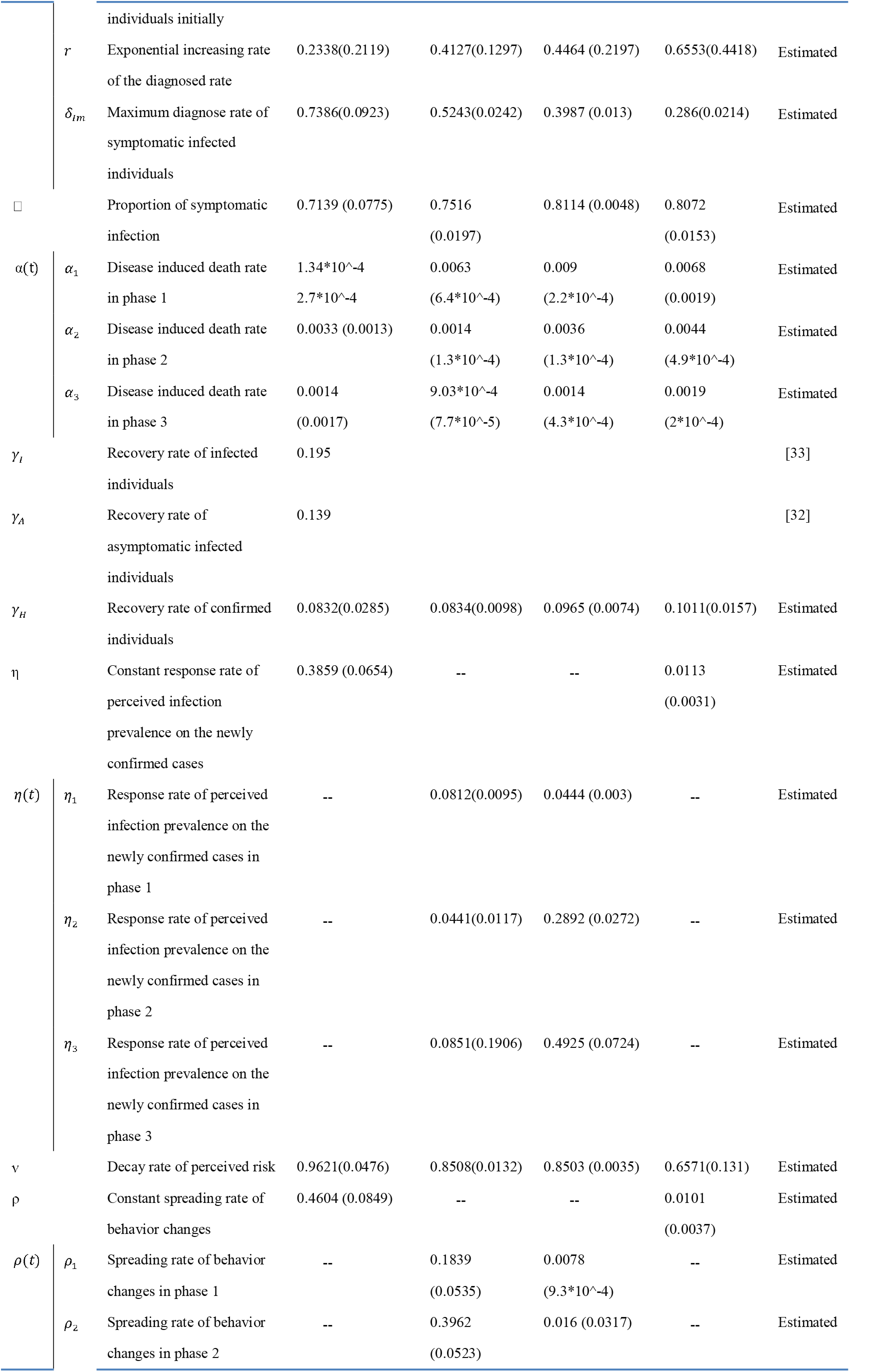
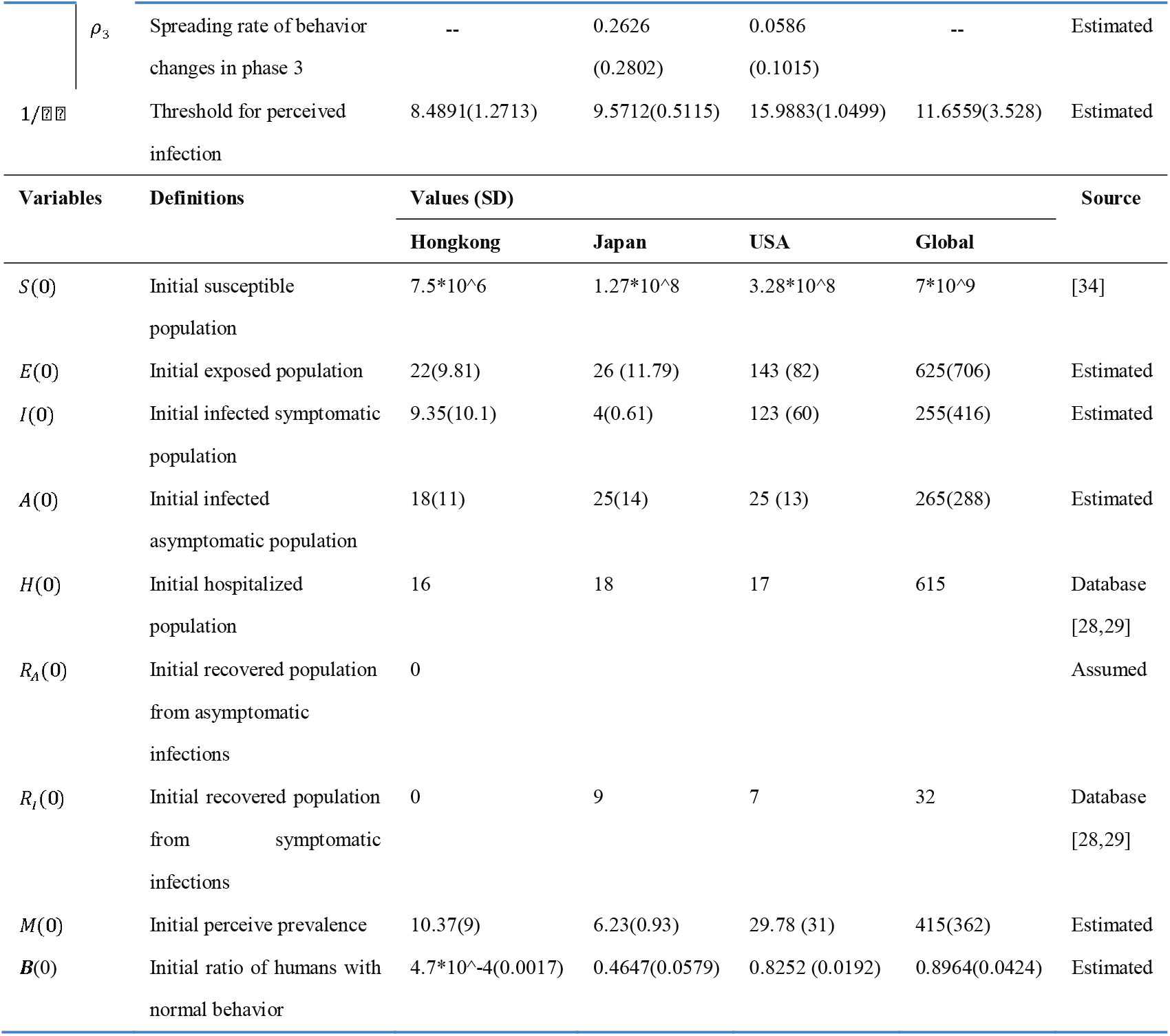
Parameter definition and estimation.

| Parameters | Definition | Value (SD) |  |  |  | Source |
| --- | --- | --- | --- | --- | --- | --- |
|  |  | Hongkong | Japan | USA | Global |  |
| $\beta_I$ | Transmission rate of symptomatic infected individuals | 1.5053(0.2508) | 1.4059(0.0521) | 2.0247(0.0571) | 1.5589 (0.1371) | Estimated |
| $\beta_A$ | Transmission rate of asymptomatic infected individuals | 0.6248(0.19) | 0.2261(0.0278) | 0.6004(0.0255) | 0.6034 (0.0512) | Estimated |
| q | Constant reduced transmission rate due to behavior changes | 0.3306(0.0702) | -- | -- | 0.3697 (0.0318) | Estimated |
| q(t) | $q_1$ Reduced transmission rate due to behavior changes in Phase 1 | -- | 0.0895(0.0580) | 0.3055 (0.01) | -- | Estimated |
| | $q_2$ Reduced transmission rate due to behavior changes in Phase 2 | -- | 0.493(0.0219) | 0.3505 (0.0084) | -- | Estimated |
| | $q_3$ Reduced transmission rate due to behavior changes in Phase 3 | -- | 0.6742(0.0189) | 0.3926 (0.0084) | -- | Estimated |
| $\sigma$ | Transmission rate of exposed individuals to the infected class | 1/5.2 | | | | [31] |
| $\delta_i(t)$ | $\delta_{i0}$ Diagnose rate of symptomatic infected | 0.0048(0.004) | 0.0081(0.0015) | 0.0079 (7.7*10<sup>-4</sup>) | 0.0084(0.001) | Estimated |

|  |  |  |  |  |  |  |  |
| --- | --- | --- | --- | --- | --- | --- | --- |
|  |  | individuals initially |  |  |  |  |  |
| | $r$ | Exponential increasing rate of the diagnosed rate | 0.2338(0.2119) | 0.4127(0.1297) | 0.4464 (0.2197) | 0.6553(0.4418) | Estimated |
| | $\delta_{Im}$ | Maximum diagnose rate of symptomatic infected individuals | 0.7386(0.0923) | 0.5243(0.0242) | 0.3987 (0.013) | 0.286(0.0214) | Estimated |
| □ | | Proportion of symptomatic infection | 0.7139 (0.0775) | 0.7516 (0.0197) | 0.8114 (0.0048) | 0.8072 (0.0153) | Estimated |
| $\alpha(t)$ | $\alpha_1$ | Disease induced death rate in phase 1 | 1.34*10<sup>-4</sup><br>2.7*10<sup>-4</sup> | 0.0063<br>(6.4*10<sup>-4</sup>) | 0.009<br>(2.2*10<sup>-4</sup>) | 0.0068<br>(0.0019) | Estimated |
| | $\alpha_2$ | Disease induced death rate in phase 2 | 0.0033 (0.0013) | 0.0014<br>(1.3*10<sup>-4</sup>) | 0.0036<br>(1.3*10<sup>-4</sup>) | 0.0044<br>(4.9*10<sup>-4</sup>) | Estimated |
| | $\alpha_3$ | Disease induced death rate in phase 3 | 0.0014<br>(0.0017) | 9.03*10<sup>-4</sup><br>(7.7*10<sup>-5</sup>) | 0.0014<br>(4.3*10<sup>-4</sup>) | 0.0019<br>(2*10<sup>-4</sup>) | Estimated |
| $\gamma_I$ | | Recovery rate of infected individuals | 0.195 | | | | [33] |
| $\gamma_A$ | | Recovery rate of asymptomatic infected individuals | 0.139 | | | | [32] |
| $\gamma_H$ | | Recovery rate of confirmed individuals | 0.0832(0.0285) | 0.0834(0.0098) | 0.0965 (0.0074) | 0.1011(0.0157) | Estimated |
| $\eta$ | | Constant response rate of perceived infection prevalence on the newly confirmed cases | 0.3859 (0.0654) | -- | -- | 0.0113 (0.0031) | Estimated |
| $\eta(t)$ | $\eta_1$ | Response rate of perceived infection prevalence on the newly confirmed cases in phase 1 | -- | 0.0812(0.0095) | 0.0444 (0.003) | -- | Estimated |
| | $\eta_2$ | Response rate of perceived infection prevalence on the newly confirmed cases in phase 2 | -- | 0.0441(0.0117) | 0.2892 (0.0272) | -- | Estimated |
| | $\eta_3$ | Response rate of perceived infection prevalence on the newly confirmed cases in phase 3 | -- | 0.0851(0.1906) | 0.4925 (0.0724) | -- | Estimated |
| $\nu$ | | Decay rate of perceived risk | 0.9621(0.0476) | 0.8508(0.0132) | 0.8503 (0.0035) | 0.6571(0.131) | Estimated |
| $\rho$ | | Constant spreading rate of behavior changes | 0.4604 (0.0849) | -- | -- | 0.0101 (0.0037) | Estimated |
| $\rho(t)$ | $\rho_1$ | Spreading rate of behavior changes in phase 1 | -- | 0.1839 (0.0535) | 0.0078 (9.3*10<sup>-4</sup>) | -- | Estimated |
| | $\rho_2$ | Spreading rate of behavior changes in phase 2 | -- | 0.3962 (0.0523) | 0.016 (0.0317) | -- | Estimated |

| $\rho_3$ | Spreading rate of behavior changes in phase 3 | -- | 0.2626<br>(0.2802) | 0.0586<br>(0.1015) | -- | Estimated |
| --- | --- | --- | --- | --- | --- | --- |
|  | Threshold for perceived infection | 8.4891(1.2713) | 9.5712(0.5115) | 15.9883(1.0499) | 11.6559(3.528) | Estimated |
| Variables | Definitions | Values (SD) |  |  |  | Source |
|  |  | Hongkong | Japan | USA | Global |  |
| $S(0)$ | Initial susceptible population | $7.5 \times 10^6$ | $1.27 \times 10^8$ | $3.28 \times 10^8$ | $7 \times 10^9$ | [34] |
| $E(0)$ | Initial exposed population | 22(9.81) | 26 (11.79) | 143 (82) | 625(706) | Estimated |
| $I(0)$ | Initial infected symptomatic population | 9.35(10.1) | 4(0.61) | 123 (60) | 255(416) | Estimated |
| $A(0)$ | Initial infected asymptomatic population | 18(11) | 25(14) | 25 (13) | 265(288) | Estimated |
| $H(0)$ | Initial hospitalized population | 16 | 18 | 17 | 615 | Database<br>[28,29] |
| $R_A(0)$ | Initial recovered population from asymptomatic infections | 0 | | | | Assumed |
| $R_I(0)$ | Initial recovered population from symptomatic infections | 0 | 9 | 7 | 32 | Database<br>[28,29] |
| $M(0)$ | Initial perceive prevalence | 10.37(9) | 6.23(0.93) | 29.78 (31) | 415(362) | Estimated |
| $B(0)$ | Initial ratio of humans with normal behavior | $4.7 \times 10^{-4}$(0.0017) | 0.4647(0.0579) | 0.8252 (0.0192) | 0.8964(0.0424) | Estimated |

### 2.2 Evolution of behavioral changes and multiple epidemic waves

Based on the model fitting results, we obtained the estimate of evolution of perceived infection *(M(t))* and the dynamics of proportion of the humans with behavioral changes (1 − *B(t))* for three regions/countries and the world, as shown in Fig. 2. It is worth noting that the estimated perceived infection and its corresponding behavioral changes behave oscillatively, in particular, for Japan and Hongkong, which might be the main factor to induce multiple epidemic waves in these two regions/countries. Taking Hongkong as an example, as the epidemic initially increased quickly, the individuals consciously or compulsively chose to change their behaviors to reduce the risk of being infected and the proportion of the population with behavioral changes quickly increased close to 100% around the time of the first epidemic peak. Then, the quickly changed behaviors induced the epidemic to decline swiftly and the daily reported cases reduced to almost zero. This in turn drove individuals to return back to normal behaviors, and consequently the susceptible population size rebounded back to a higher level, which induced another epidemic wave. This feedback loop, epidemic increasing → behavior changing → epidemic declining → behavior changing back → epidemic resurging, could repeat multiple times to drive multiple COVID-19 epidemic waves as observed in Hongkong or Japan (Figs. 1 and 2). From Fig. 1, we can see that the proposed model by integrating behavioral change dynamics and the disease transmission dynamics could capture the observed multiple waves of COVID-19 epidemics very well.

**Fig. 2.**
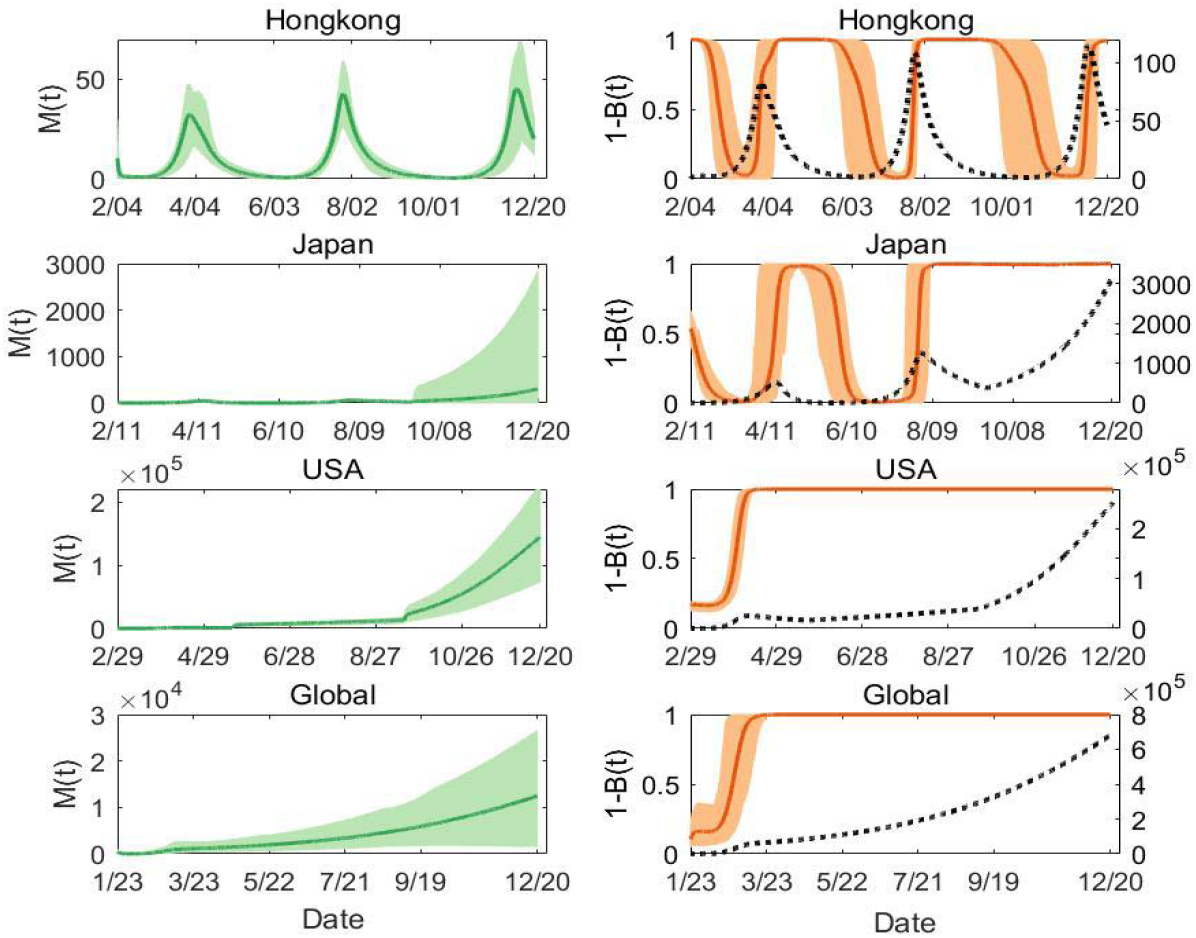
Estimated behavioral dynamics: 1 −*B*(*t*) and *M* (*t*). The solid curves are the estimated dynamic curves with the shadow areas as the corresponding 95% confidence intervals based on the bootstrap method. The black dash curves are the estimated daily reported cases from Model (1).

Note that we also observed the third epidemic wave in Japan that lasted longer and higher than the first two waves, although most people chose to change their behaviors during the third wave. Similar phenomenon was also observed in USA and the global COVID-19 epidemics (Fig. 2). Then, the interesting question is why the behavioral change did not significantly induce the epidemic to decline in the later waves. To address this question, we further examined the evolution of key parameters related to behavioral changes. The estimates of behavior-related dynamic parameters, the reduced transmission rate due to alternative behaviors (q), the response rate of perceived infection prevalence (η), and the spreading rate of behavior changes (ρ) for Japan and USA are shown in Fig. 3. We can see that the parameter q(t), the reduced transmission rate due to alternative behaviors exhibited an increasing trend during the multiple waves for USA and Japan. This suggests that the effect of behavioral changes in terms of reduction in transmission rate significantly decreased in later waves either due to waning of adherence to the NPIs or pandemic fatigue (i.e., fatigue of risk awareness of self-protection). Therefore, not only the proportion of the population, but also the effectiveness of alternative behaviors to reduce the risk of being infected is critical to prevent the epidemic from continuously surging.

**Fig 3.**
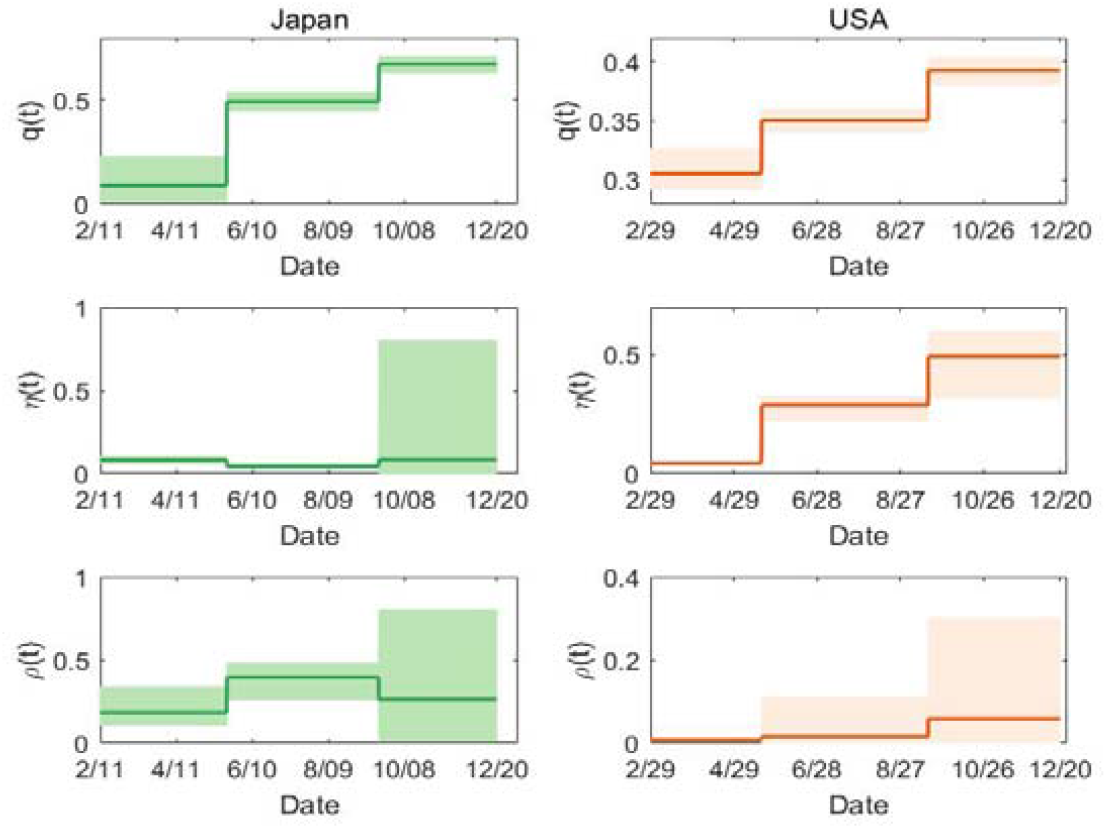
Evolution of the estimated values of the three key parameters related to behavioral changes in Japan and USA.

### 2.3 Effect of behavioral changes on controlling multiple COVID-19 epidemic waves

We further evaluated whether the subsequent epidemic waves could be avoided if the effectiveness of behavioral changes could be enhanced after the first wave. If the adherence of NPIs and awareness of self-protection were enhanced so that the reduced transmission rate due to the alternative behaviors (q) during the second and third waves was the same as that during the first wave, the simulated daily new cases in USA and Japan were shown in Fig. 4(A-B), from which we can see that the magnitude of second and third waves could be significantly reduced or completely avoided. This suggests that it is very important to promote and maintain the high awareness of self-protection and well adherence to the NPIs in order to avoid subsequent epidemic waves. Similarly, by controlling other behavioral change parameters, (*η, ρ, m, v*), it could also mitigate COVID-19 epidemics, which is demonstrated for the case of Hongkong in Fig. 4(C-F). That is, the peak magnitude of subsequent waves could be reduced by accelerating the risk awareness, spreading of behavioral changes and increasing the sensitivity of individuals to perceived infection (i.e., increasing the values of *(η, ρ,m))* as well as prolonging the period of higher risk awareness (reducing the value of *v*).

**Fig. 4.**
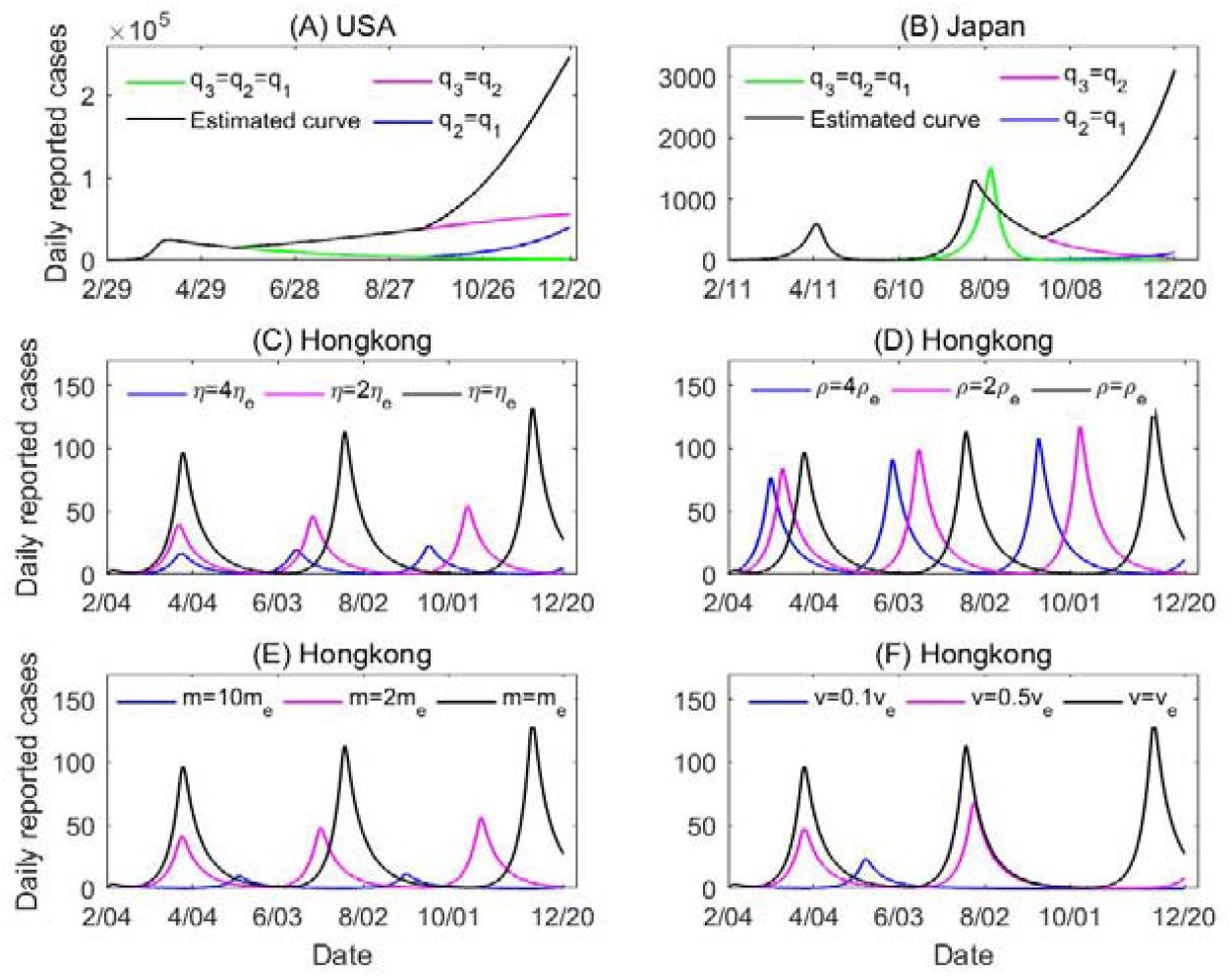
Simulated daily reported cases based on Model (1) with different assumptions of behavioral dynamic parameters. (A)-(B) Daily reported cases in USA and Japan under different scenarios by decreasing the values of *q* in Phases 2 and 3. (C)-(F) Daily reported cases in Hongkong by varying the four parameters related to behavioral change dynamics. The subscript ‘e’ indicates the estimated value from the observed data. The values of all other parameters were set as those listed in Table 1.

### 2.4 Relaxation of NPIs under vaccination

The COVID-19 vaccines started to be available in some countries from December 2020. With vaccination, people’s perceived risk of infection might be changed and this may affect the behavioral changes. To investigate the evolution of behavioral changes under vaccination and the trade-off effect between relaxation of NPIs and vaccination on COVID-19 epidemics, we extended Model (1) to include the effect of vaccination, see Model (2) in sections of Methods. We re-estimated the parameters related to behavioral changes, (*q, ρ, η,m,v*) and other vaccination-related parameters by fitting Model (2) to the observed epidemic data (daily new cases) and daily vaccinated population data between Dec 21st, 2020 to Feb 14th, 2021 in the USA where the COVID-19 vaccination was implemented during this period. We fixed other model parameters as those in Model (1) before vaccination started. The model fitting results are shown in SI Fig. 4. The updated estimates of behavioral change-related parameters *q*_*4*_, *ρ* _*4*_, *η*_*4*_,*m*_*A*_, *and v*_*4*_ in the fourth phase (i.e., the phase with vaccination) and the estimated values of vaccination rates *μ* _0_,*r* _*μ*_, *and μ*_*b*_. are listed in Table 2. From Table 1 and Table 2, we can see that the reduced transmission rate due to alternative behaviors *q* is less than that in the previous two phases (i.e., q_4_ < *q*_*2*_ *< q*_*3*_), which indicates the enhanced implementation of NPIs and/or the greater adherence to NPIs during the 4^th^ phase. We can also see from SI Fig. 4(C) that the vaccination coverage (received two doses of vaccines) only reached around 5% by Feb 14, 2020, which was still far from enough.

**Table 2.**
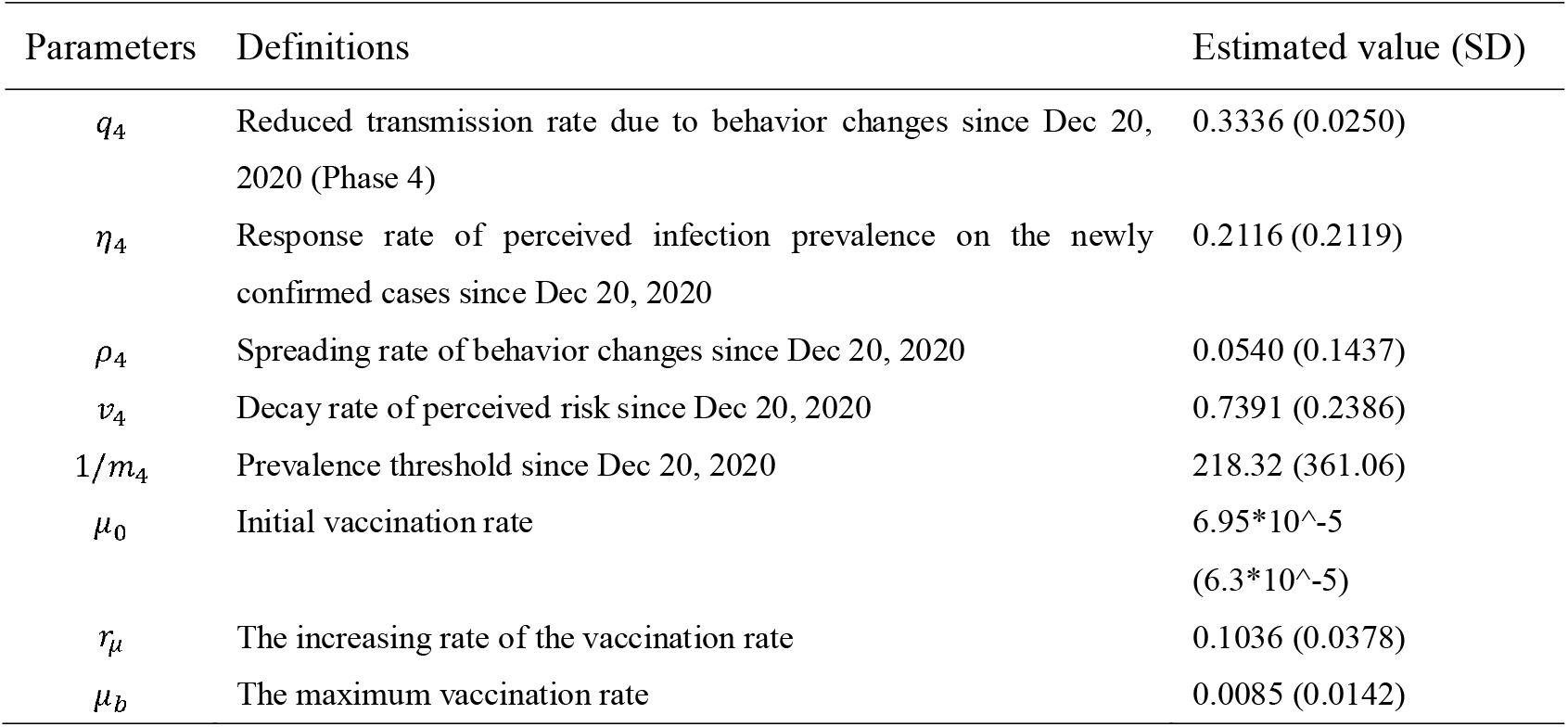
Definitions and estimated values of additional parameters in the vaccination model (2) for USA.

Note that the number of daily reported cases in USA has been declining since Dec 20th, 2020. To identify which factor mainly contributed to the declining trend, we did simulations for the epidemic from December 20 to June 30, 2021 using the established Model (2) by considering two scenarios of the reduced transmission rate due to alternative behaviors q: one as the estimated values for the 4^th^ phase and another as returning to the higher level of Phase 3, under the condition with and without vaccination (the vaccination rate μ_0_ was set as the estimated values for the 4^th^ phase). The simulated results for the four cases are shown in Fig. 5(A), from which we can see that the epidemic curves with and without vaccination are very close to each other during the period between Dec 20th, 2020 to Feb 14th, 2021, but the difference in epidemic curves between different values of q is much bigger. This indicates that the key factor for significant decline of observed daily cases during this period is more likely due to the altered behavioral changes, instead of vaccination effect. While, as the vaccination persistently continues, it starts to play its role in mitigating the epidemic, which could be seen by a big divergence between the two epidemic curves with and without vaccination in later times.

**Fig. 5.**
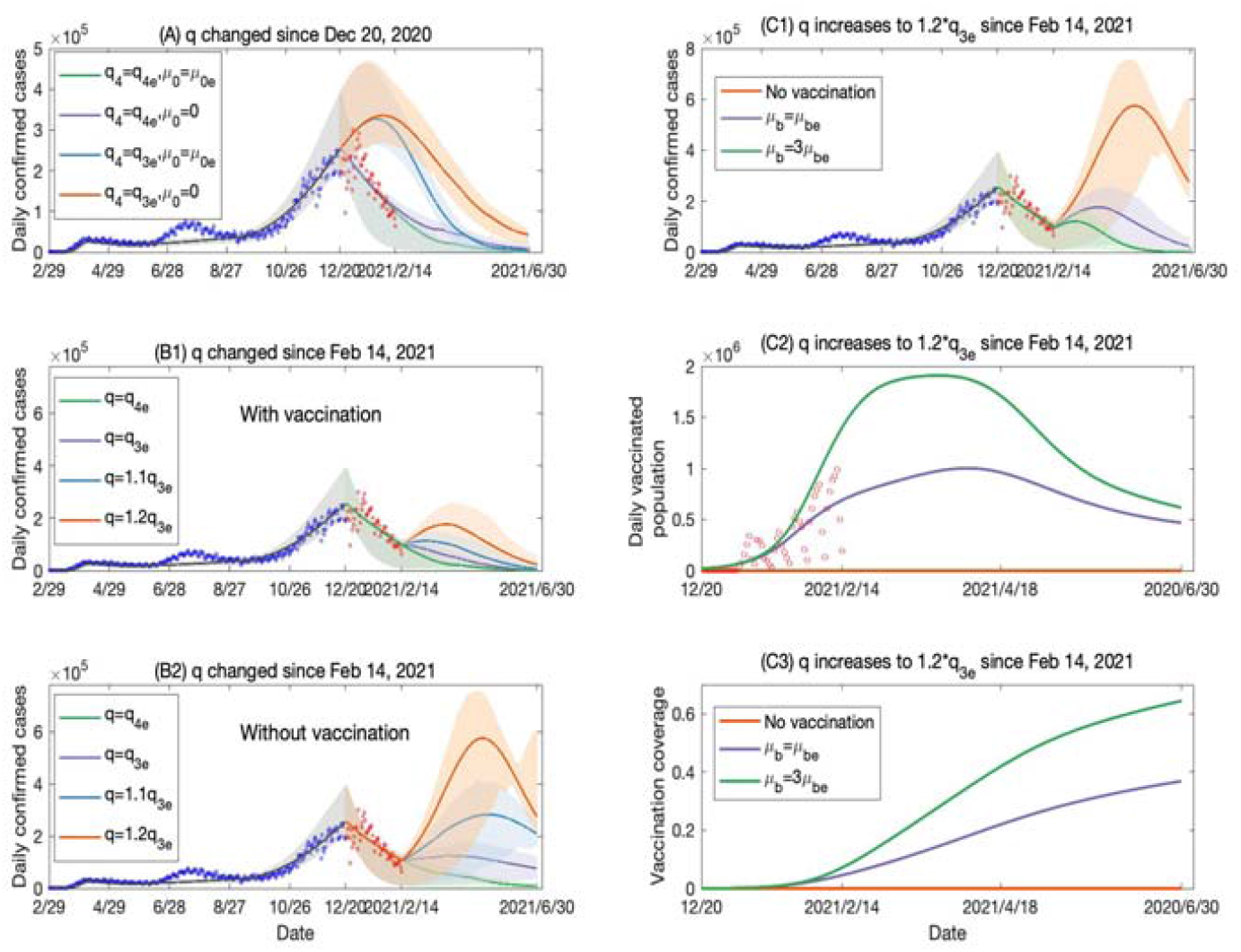
Simulation and prediction results from the vaccination model (2) for USA. (A) Two scenarios of the reduced transmission rate due to alternative behaviors (*q*): one as the estimated values for the 4^th^ phase 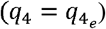 and another as returning to the higher level of Phase 3 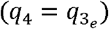, under the condition with vaccination (the vaccination rate p_0_ was set as the estimated values for the 4^th^ phase, μ_0_ = μ_0e_) and without vaccination (*μ*_0_ = 0). (B1)-(B2) *q* increased since Feb 14, 2021, where the vaccination rate was set as the estimated value from the data (B1) and no vaccination was used (B2). (C1)-(C3) *q* increased to 20% higher than that in Phase 3 since Feb 14, 2021 and the different vaccination rates were used. The vaccination coverage can reach around 40% till the end of June 2021 under the estimated vaccination rate from the current data, while the coverage could increase to over 60% by tripling the maximum vaccination rate *μ*_*b*_. The subscript ‘e’ indicating the estimated value from the data.

It is expected that the human behaviors may be likely to change to less adherence to NPIs with widespread availability of COVID-19 vaccines, which would inevitably induce a higher level of the reduced transmission rate due to alternative behaviors (q). We performed simulations to predict the COVID-19 epidemic trend from Feb 14th to June 30^th^, 2021 (Phase 5) using the established Model (2) by assuming *q* to increase from q_4_, q_3_, and 10% to 20% higher than q_3_. The two cases with and without vaccination (the vaccination rate u_0_ was set as the estimated value for the 4^th^ phase) were simulated and the results are shown in Fig. 5(B1) and (B2), respectively. The predicted results show that a higher value of *q* due to relaxation of NPIs could induce another big epidemic wave in USA during Phase 5 without vaccination (Fig. 5(B2)); but fortunately, continued vaccination could flatten the new wave significantly (Fig. 5(B1)). To further evaluate the effect of accelerated vaccination during Phase 5, we also simulated the scenario that the maximum vaccination rate would be tripled during Phase 5 under the worst condition of reduced transmission rate due to alternative behaviors, *q =* 1.2 *q*_*3*_. The results are shown in Fig. 5(C1)-(C3), indicating that the accelerated vaccination could effectively flatten or even avoid the subsequent epidemic waves due to relaxation of NPIs.

## Discussion and conclusion

Multiple waves were clearly observed from many local and nationwide COVID-19 epidemic data since early 2020 (SI Fig. 1). It is critical to understand the underlying mechanism and identify the main drivers for multiple epidemic waves in order to prevent future COVID-19 waves and outbreaks [24-26]. In this study, we proposed a multi-scale model by linking the behavioral change dynamics to the disease transmission dynamics [18-20]. We explicitly modeled the perceived infection *(M(t))* and the proportion of individuals who have altered their behaviors (1 − *B(t))*, with key behavioral change parameters such as the sensitivity to the infection risk, the speed of raising the risk awareness, the persistence to maintain the risk awareness, and the spreading rate of behavioral changes among individuals. The effect of vaccination was also considered in the model. We fitted the proposed model to the COVID-19 epidemic data with multiple waves from different regions/countries, from which the extended SEIR model, coupled with the model of behavioral change dynamics, could fit the data well and flexibly capture the asymmetric dynamics with multiple waves by considering pandemic fatigue (waning of adherence to the NPIs). The game theory based on the feedback loop between behavioral changes and COVID-19 transmission dynamics could be used to explain the observed multiple epidemic waves. Based on the developed multi-scale model with the estimated behavioral dynamic parameters and vaccination-related parameters, we also investigated the interplays among vaccine uptake, behavioral change and relaxation of NPIs as well as their effects on future COVID-19 epidemics.

The main modeling results revealed that the behavioral change dynamics, driven by the perceived COVID-19 epidemic and its effect on adherence of NPIs, played an essential role in inducing multiple epidemic waves. Our findings also suggest that continuous increases in reported new cases and persistent promotion of NPIs could result in almost whole population changing their behaviors and maintaining the altered behaviors (Fig. 2), however, the effect of behavioral changes on mitigating the epidemic was weakened (Fig. 3), presumably because of pandemic fatigue and lower adherence to the NPIs. This could induce even higher subsequent epidemic waves. Note that the three behavioral dynamic parameters, the sensitivity to the infection risk (*m*), persistence of maintaining the risk awareness *(v)*, and speed of raising risk awareness (*η*) play more important roles in mitigating the COVID-19 epidemics and avoiding the subsequent waves (Fig.4).

Given the availability of COVID-19 vaccines at the end of 2020, it is expected to gradually lift the NPIs to restore to normal life in 2021. However, our modeling results show that it should be cautious to avoid relaxing NPIs prematurely. During the early stage of vaccination, relaxation of NPIs may induce another big wave of COVID-19 epidemic. With continuous vaccination when a significant proportion of the population are vaccinated, the vaccine effect could counteract the relaxing of NPIs to avoid a new epidemic wave as expected. Thus, accelerated vaccination could allow to lift NPIs and restore normal life earlier. But the interplay between vaccine uptake and relaxation of NPIs should be carefully evaluated and the caution should be taken before relaxing NPIs.

Note that we ignored the mutations of SARS-CoV-2 in our model, the high transmission rate of the mutated strains may raise risk awareness and consequently induce a new round of behavioral changes. Complicated behavioral dynamics, driven by multiple factors including efficacy of the COVID-19 vaccines, transmission properties of the mutated strains, relaxation of NPIs among others, may need to be considered in the future work. It is not trivial to estimate the behavioral dynamic parameters based on the observed epidemic data only, especially the identifiability of nonlinear differential equation models is a fundamental and challenging problem [27]. Collection and use of behavioral data during the epidemic period could fill the gap to further refine the proposed model in the future.

## Methods

### Data

The COVID-19 epidemic data were obtained from the Johns Hopkins University Center for Systems Science and Engineering (JHU CCSE) and the data are available on the Github and the Humanitarian Data Exchange [28,29]. We used the data of daily COVID-19 confirmed cases and accumulative death cases in Hongkong, Japan, USA, and the world (including 273 countries or regions), as shown in SI Fig.1. We assumed that the local community transmission begun when there were continuously reported cases for 5 days. Consequently, the data collected in Hongkong, Japan, USA, and the whole world started on February 4th, 11th, 29th, and January 23rd, 2020 respectively. As we can see from the epidemic data, Hongkong, Japan and USA have already experienced multiple epidemic waves during 2020, and the last wave was much more serious with a much higher peak in Japan and USA than that of the first two waves. We obtained the data of the daily vaccinated population in USA (who received two doses) from the US Centers for Disease Control and Prevention (CDC) [30]. The data were released and analyzed anonymously [28-30].

### The multi-scale dynamic models

Individuals may reduce their susceptibility or infectivity by changing behaviors in response to the growing epidemic. Reduction of contact rate or transmission probability can be achieved by taking self-protective actions such as keeping social distancing, wearing masks, and washing hands. To model the behavioral changes triggered by the scare of infection, we propose a co-evolution model linking the disease transmission dynamics to the behavioral change dynamics (SI Fig. 2).

The SEIR model is modified to an SEIAHR model for disease transmission dynamics by considering symptomatic/asymptomatic infected individuals. The total population *N* is divided into susceptible individuals (*S*), exposed individuals *(E)*, infectives with symptoms *(I)*, asymptomatic infectives (*A*), hospitalized individuals *(H)*, recovered individuals (R_*I*_) and asymptomatic recovered individuals *(R*_*A*_). The perceived risk of infection contributes to the evolution of human behavioral changes in response to the scare of being infected. Then the perceived infection (called information index, denoted by M) or information spreading process is assumed to be related to the number of new confirmed cases and faded through an exponentially fading memory model. The dynamics of behavioral changes can be described by the imitation dynamics, which is a learning process. Expected payoffs of two different behavioral strategies (say, normal or altered behavior) are compared with each other, and individuals may change their strategy when they become aware that their payoff can be increased by adopting an alternative behavior. Let *B* be the fractions of population who perform no behavioral changes. Then, the model can be written as,

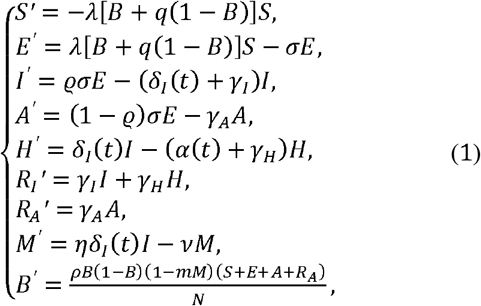

where 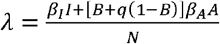. Parameter 0 ≤ q ≤ 1 is the reduced transmission rate due to the altered behavior in response to the risk of infection. If q = 0, the transmission rate is reduced to 0 because of the altered behavior; if q = 1, the altered behavior does not change the transmission rate. Parameter 1/*σ* is the incubation period and *Q* is the probability of suffering symptoms after being infected. *δ*_*I*_(t) is the detection rate of symptomatically infected individuals and *α (t)* is the disease-induced death rate. Parameters (*γ*_*I*_, *γ*_*A*_,*γ*_*H*_) are the recovery rates of symptomatic infected, asymptomatic infected and hospitalized individuals, respectively. Parameter *η* is the response rate of perceived infection because of the number of newly confirmed cases, *v* is the decay rate of the perceived risk, and *m* is the sensitivity of individuals to perceived infection, where a higher value of *m* indicates that individuals perceive the disease as more dangerous. Parameter *ρ* represents the speed of the imitation process with respect to the disease transmission dynamics. The definitions and values of other parameters are given in Table 1.

Considering the continuous improvement of testing capacity, the diagnosis rate is set to be an increasing function of time t with the following form [35]:

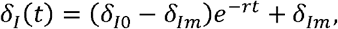

where δ_I0_ is the initial diagnosis rate while δ_Im_ is the maximum diagnosis rate, and r is the corresponding exponential increasing rate. As reported in several studies [36,37], the COVID-19 fatality rate is varying overtime. Hence, the disease-induced death rate *α* is set to be a piecewise function of time t with three phases:

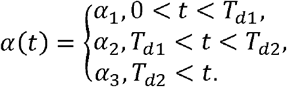

Here, we assumed that T_dl_ = 150,100,80,80 (day) and T_d2_ = 250,220,200,200 (day) in Hongkong, Japan, USA, and global area, respectively. Similarly, to represent the behavioral change-epidemic feedback loop for multiple waves in Japan and USA (see SI Fig.1), we chose piecewise smooth functions for behavioral dynamics-related parameters (*q, ρ, η*), see details in SI. Note that, although Hongkong experienced three similar epidemic waves till December, 2020, we set the behavioral dynamics-related parameters (*q, ρ, η*) as positive constants during the time period considered in this study. The global epidemic of COVID-19 only experienced one epidemic wave (still in the increasing phase) till Dec 20th, 2020, we then set these parameters as constants for the global epidemic.

Considering a continuous vaccination regime, we can extend Model (1) to the following system by considering vaccine effect,

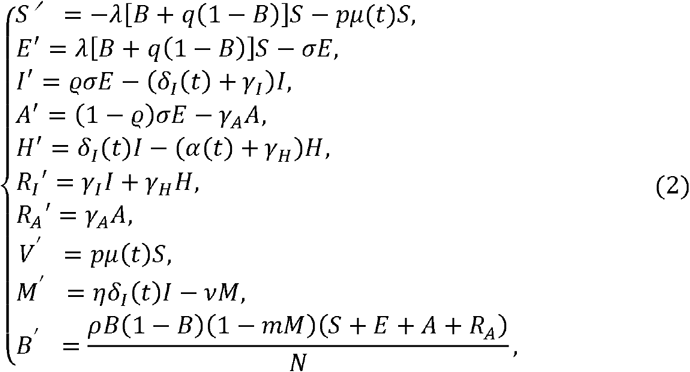

where *μ (t)* is the time-dependent vaccination rate, as it should be small initially since the availability of the number of vaccine doses was limited at the beginning in December 2020, and then exponentially increased as the production of COVID-19 vaccines was accelerated, and finally it could be plateaued to a constant level depending on the daily vaccination capacity. Thus, we model *μ (t)* as a logistic increasing function of time *t* with the following form:

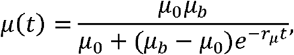

where *μ*_0_ (*μ*_*b*_) is the initial (maximum) vaccination rate, *r*_*μ*_ is the exponential increasing rate of the vaccination. The vaccination efficacy is denoted by *p* with *p =* 95% in the USA [38,39] and the effectively vaccinated population is assumed to be immune to COVID-19 and moved to the class *V* (the effectively vaccinated compartment). Consequently, the time-dependent vaccination coverage can be defined as:

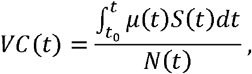

where *t*_0_ is the starting time of vaccination, which was set as Dec 20th, 2020 for USA.

### Basic reproduction number

When all individuals are adopting the normal behavior, i.e., B=1, the basic reproduction number can be obtained as

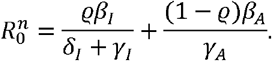

When all individuals are adopting the altered behaviors, i.e., B=0, the basic reproduction number can be derived as

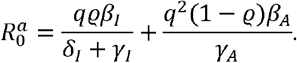

When considering the co-evolution of behavioral changes and the disease transmission dynamics (B ≠ 0 and B ≠ 1), the proportion of individuals adopting altered behaviors and the number of susceptible individuals are time-varying, under which we could obtain the time-varying reproduction number *R(t)*, referring to as the effective reproduction number,

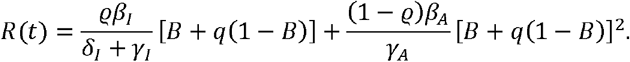

This is a combination of two basic reproduction numbers.

### Model fitting and parameter estimation methods

We fixed some parameters in Model (1) including the incubation period (1/*σ*), recovery rate of asymptomatic infections (*γ*_*A*_) and symptomatic infections (*γ*_*I*_), based on the existing literature as listed in Table 1. The initial total population was fixed as the whole population in the corresponding regions or countries, and the initial hospitalized and recovered populations were obtained from the database (Table 1) [28,29]. We generate 500 bootstrap samples of the time series data of daily reported cases and accumulative death cases based on a Poisson distribution (the mean of the Poisson distribution was assumed as the observed counts). We first fitted Model (1) to the 500 bootstrap samples of daily reported cases and accumulative death cases using the least squared (LS) method. We then fitted Model (2) to the 500 bootstrap samples of daily reported cases and the daily vaccinated population in USA by fixing the parameters in Model (1), except the behavior change related and vaccination related parameters. In more details, we used the least square method with a *priori* distribution for each parameter to fit the model to the data using the software of Matlab, where the ODE system is solved by the “ODE45” function while the “fmincon” function is used to search the optimal solutions of the objective function.

## Data Availability

All the data used in this study is public available, and the related link is provided in the manuscript.

## Author contributions

Conceptualization, YX, ST; validation and simulation, BT, WZ, XW, ST; data curation, BT, WZ, ST; writing—original draft preparation, BT, YX; writing—review and editing, HW, YX; All authors have read and agreed to the published version of the manuscript.

