## Supplementary Information for "Controlling multiple COVID-19 epidemic waves: an insight from a multi-scale model linking the behavior change dynamics to the disease transmission dynamics"

We provide the details for the COVID-19 transmission dynamic model and the behavioral change dynamic model first, and then integrate the two models into a co-evolution multi-scale model.

**Model assumptions**

The SEIR model is modified to an SEIAHR model to describe the transmission dynamics of COVID-19 by considering that infective individuals may suffer symptoms or experience no symptoms. In details, the total population N is divided into susceptible individuals ($S$), exposed individuals ($E$), infectives with symptoms ($I$), asymptomatic infectives ($A$), hospitalized individuals ($H$) and symptomatic recovered individuals ($R_{I}$) and asymptomatic recovered individuals ($R_{A}$). During an outbreak of an emerging infectious disease, behavioral changes likely occur in order to reduce the infection risk. Note that, though only susceptible individuals are exposed to the risk of COVID-19 infection, asymptomatic infected individuals and recovered individuals who did not suffer symptoms have no awareness of their infection, and thus would behave similarly as susceptible individuals. Thus, in this study, we assumed that the self-protection behaviors can be adopted by not only susceptible individuals, but also asymptomatic infectives and recovered individuals who did not experience symptoms. In contrast, we assumed that all symptomatic infected individuals would take the same actions (i.e., no behavioral changes). Thus, in our model, we assumed that individuals suffering no symptoms (S, E, A, $R_{A}$) are supposed to be able to reduce their susceptibility or infectivity by changing behaviors in response to the COVID-19 outbreak. Thus, reduction of contact rates or transmission probability (i.e., transmission rate) can be achieved by adopting self-protective behaviors such as limiting travels, keeping social distancing, wearing masks, washing hands, etc.

Basically, an individual’s behavior change is assumed to be driven by the evaluation and comparison of payoffs between the two behaviors. With the cost-benefit consideration, individuals are supposed to change their behaviors when they realized that the other behavior is more beneficial with low cost. The integrated co-evolution model is the result of coupled two dynamic processes: the disease transmission process and the behavioral change process, as illustrated in SI Fig. 2. The two processes were modelled in two different time scales since the contacts related to the transmission of virus are physical person-to-person interactions, which is less frequently than the contacts related to the spread of information, which could be accessed by telephone, media and online, etc. Thus, we introduce two time-units: $t$ as the time unit of disease transmission and $\tau$ as the time unit of behavioral changes, and assume that $t=\alpha\tau$, where $\alpha$ is a scaling parameter.

**Disease transmission dynamics**

With the above assumptions, the susceptible, exposed, asymptomatic infected and asymptomatic recovered individuals are divided into two subclasses: individuals adopting the normal behavior ($S_{n},E_{n},{A_{n},R}_{A_{n}}$) and individuals adopting the altered behavior ($S_{a},E_{a},{A_{a},R}_{A_{a}}$). Then the disease transmission process model is given as follows:

$\left\{ \begin{aligned} &{S_{n}}^{'}(t)=-\lambda S_{n}, \\ &{S_{a}}^{'}(t)=-q\lambda S_{a}, \\ &{E_{n}}^{'}(t)=\lambda S_{n}-\sigma E_{n}, \\ &{E_{a}}^{'}(t)=q\lambda S_{a}-\sigma E_{a}, \\ &{I(t)}^{'}=\varrho\sigma E_{n}+\varrho\sigma E_{a}-\left( \delta_{I}\left( t \right)+\gamma_{I} \right)I, \\ &{A_{n}\left( t \right)}^{'}=\left( 1-\varrho\right)\sigma E_{n}-\gamma_{A}A_{n}, \\ &{A_{a}\left( t \right)}^{'}=\left( 1-\varrho\right)\sigma E_{a}-\gamma_{A}A_{a}, \\ &{H\left( t \right)}^{'}=\delta_{I}\left( t \right)I-\left( \alpha\left( t \right)+\gamma_{H} \right)H, \\ &{R_{I}\left( t \right)}^{'}=\gamma_{I}I+\gamma_{H}H, \\ &{R_{A_{n}}\left( t \right)}^{'}=\gamma_{A}A_{n}, \\ &{R_{A_{a}}\left( t \right)}^{'}=\gamma_{A}A_{a}, \end{aligned} \right.$	(S1)

where $\lambda=\left( \beta_{I}I+\beta_{A}A_{n}+q\beta_{A}A_{a} \right)/N$ is the force of infection, which is composed of three parts: the infection by symptomatic infected individuals ($\beta_{I}I/N$), the infection by asymptomatic infected individuals adopting the normal behavior ($\beta_{A}A_{n}/N$), and the infection by asymptomatic infected individuals adopting the altered behavior ($q\beta_{A}A_{a}/N$). ${(\beta}_{I},\beta_{A})$ are the transmission rates of symptomatic infected and asymptomatic infected individuals, respectively. As behavior changed, the infectivity of asymptomatic infected individuals and the susceptibility of susceptible individuals could be reduced, and parameter $q$($0\leq q\leq1$) is the corresponding reduced transmission rate due to alternative behaviors. $1/\sigma$ is the incubation period and $\varrho$ is the probability with symptoms after being infected.$\delta_{I}\left( t \right)$ is the time-varying diagnosis rate of symptomatic infected individuals and $\alpha\left( t \right)$ is the time-dependent disease-induced death rate.${(\gamma}_{I}, \gamma_{A},\gamma_{H})$ are the recovery rates of symptomatic infected, asymptomatic infected and confirmed individuals, respectively.

**Behavioral change dynamics**

Models by integrating disease transmission dynamics and behavioral change dynamics have been proposed to study the spread of infectious diseases, including COVID-19 [15-20]. The modeling framework and concept of game theory were used to explain the behavioral change dynamics during an outbreak of infectious diseases and their effect on the disease epidemics. Here we review and use these modeling framework and concepts to develop our multi-scale models for COVID-19 epidemics.

**Perceived infection:** The perceived risk of infection contributes to the evolution of human behavioral changes in response to the scare of being infected. Then the perceived infection (or information index) is assumed to be stimulated by the number of newly confirmed cases and faded through an exponentially memory fading mechanism. Thus, we have the following equation:

${M(t)}^{'}=\eta\delta_{I}\left( t \right)I-\nu M,$	(S2)

where $M(t)$ is an information variable governing the signal available to individuals as a function of daily reported cases, $\eta$ is the response rate of perceived infection (or information index) on the number of newly confirmed cases, and $\nu$ is the decay rate of the perceived infection (or information index).

**Payoffs:** The expected payoffs to adopt two different behaviors: normal behavior and altered behavior, which can be recognized as the negative of the costs, are assumed to be linearly dependent on the perceived infection. In addition to the cost for the risk of infection (which is assumed to be higher for normal behavior), individuals adopting altered behavior pay an extra constant cost. Thus, the payoffs with the normal behavior and the altered behavior are respectively:

$P_{n}\left( t \right)=-m_{n}M\left( t \right), P_{a}\left( t \right)=-k-m_{a}M\left( t \right),$ (S3)

with $m_{n}>m_{a}$, where $m_{n}$ and $m_{a}$ are parameters related to the risk of infection by adopting two different strategies and $k$ is the extra cost of changing behaviors.

**Imitation process:** The dynamics of behavioral changes can be described by the imitation dynamics, which is a learning process. Expected payoffs of two different behavioral strategies are compared with each other and individuals may change strategy when they become aware that their payoff can be increased if adopting another behavior. Denote $\Delta P=P_{n}-P_{a}$, then the imitation dynamics in a two-strategy game can be described by

$x^{'}=\vartheta x\left( 1-x \right)\varphi\Delta P=\omega x\left( 1-x \right)\Delta P,$(S4)

where $x$ and $1-x$ are the fractions of population performing the two strategies. $\vartheta$ is the rate at which individuals communicate with each other and $\varphi$ is the probability of changing decision. Then the sign of $x^{'}$ is determined by $\Delta P$, illustrating the changing direction of behaviors (from normal to altered or from altered to normal).

Note that the imitation process has no concern with the transition among epidemiological classes, but only drives behavior changing. Hence encounters between individuals in the imitation process would only result in migration between ${(S}_{n}$and$S_{a})$,${(E}_{n}$and $E_{a})$,${(A}_{n}$and $A_{a})$, and${(R}_{A_{n}}$and $R_{A_{a}})$. In particular, when susceptible individuals with the normal behavior ($S_{n}$) compare their payoff with the payoff of those adopting the altered behavior ($S_{a}, E_{a}, A_{a},R_{A_{a}}$), and find that $\Delta P<0$, then $S_{n}$ will mitigate to $S_{a}$, and vise versa. The migration between ${(E}_{n}$and $E_{a}$),${(A}_{n}$and $A_{a})$, and ($R_{A_{n}}$and $R_{A_{a}})$are similar. Thus, we can model the imitation dynamics in the population with two alternative behaviors as follows:

$\left\{ \begin{aligned} &{S_{n}}^{'}\left( \tau\right)=\frac{\omega S_{a}\left( S_{n}+E_{n}+A_{n}+R_{n} \right)\Delta PH\left( \Delta P \right)}{N}+\frac{\omega S_{n}\left( S_{a}+E_{a}+A_{a}+R_{a} \right)\Delta PH\left( -\Delta P \right)}{N}, \\ &{S_{a}}^{'}\left( \tau\right)=-\frac{\omega S_{a}\left( S_{n}+E_{n}+A_{n}+R_{n} \right)\Delta PH\left( \Delta P \right)}{N}-\frac{\omega S_{n}\left( S_{a}+E_{a}+A_{a}+R_{a} \right)\Delta PH\left( -\Delta P \right)}{N}, \\ &{E_{n}}^{'}\left( \tau\right)=\frac{\omega E_{a}\left( S_{n}+E_{n}+A_{n}+R_{n} \right)\Delta PH\left( \Delta P \right)}{N}+\frac{\omega E_{n}\left( S_{a}+E_{a}+A_{a}+R_{a} \right)\Delta PH\left( -\Delta P \right)}{N}, \\ &{E_{a}}^{'}\left( \tau\right)=-\frac{\omega E_{a}\left( S_{n}+E_{n}+A_{n}+R_{n} \right)\Delta PH\left( \Delta P \right)}{N}-\frac{\omega E_{n}\left( S_{a}+E_{a}+A_{a}+R_{a} \right)\Delta PH\left( -\Delta P \right)}{N}, \\ &{A_{n}\left( \tau\right)}^{'}=\frac{\omega A_{a}\left( S_{n}+E_{n}+A_{n}+R_{n} \right)\Delta PH(\Delta P)}{N}+\frac{\omega A_{n}\left( S_{a}+E_{a}+A_{a}+R_{a} \right)\Delta PH(-\Delta P)}{N}, \\ &{A_{a}\left( \tau\right)}^{'}=-\frac{\omega A_{a}\left( S_{n}+E_{n}+A_{n}+R_{n} \right)\Delta PH\left( \Delta P \right)}{N}-\frac{\omega A_{n}\left( S_{a}+E_{a}+A_{a}+R_{a} \right)\Delta PH\left( -\Delta P \right)}{N}, \\ &{R_{n}\left( \tau\right)}^{'}=\frac{\omega R_{a}\left( S_{n}+E_{n}+A_{n}+R_{n} \right)\Delta PH\left( \Delta P \right)}{N}+\frac{\omega R_{n}\left( S_{a}+E_{a}+A_{a}+R_{a} \right)\Delta PH\left( -\Delta P \right)}{N}, \\ &{R_{a}\left( \tau\right)}^{'}=-\frac{\omega R_{a}\left( S_{n}+E_{n}+A_{n}+R_{n} \right)\Delta PH\left( \Delta P \right)}{N}-\frac{\omega R_{n}\left( S_{a}+E_{a}+A_{a}+R_{a} \right)\Delta PH\left( -\Delta P \right)}{N}. \end{aligned} \right.$	(S5)

**Co-evolution model by coupling two dynamic processes**

Coupling the transmission model (S1) and the imitation model (S5), then we have,

$\left\{ \begin{aligned} &{S_{n}}^{'}(t)=-\lambda S_{n}+\frac{1}{\alpha}[\frac{\omega S_{a}\left( S_{n}+E_{n}+A_{n}+R_{n} \right)\Delta PH(\Delta P)}{N}+\frac{\omega S_{n}\left( S_{a}+E_{a}+A_{a}+R_{a} \right)\Delta PH(-\Delta P)}{N}], \\ &{S_{a}}^{'}(t)=-q\lambda S_{a}-\frac{1}{\alpha}[\frac{\omega S_{a}\left( S_{n}+E_{n}+A_{n}+R_{n} \right)\Delta PH\left( \Delta P \right)}{N}+\frac{\omega S_{n}\left( S_{a}+E_{a}+A_{a}+R_{a} \right)\Delta PH(-\Delta P)}{N}], \\ &{E_{n}}^{'}(t)=\lambda S_{n}-\sigma E_{n}+\frac{1}{\alpha}[\frac{\omega E_{a}\left( S_{n}+E_{n}+A_{n}+R_{n} \right)\Delta PH(\Delta P)}{N}+\frac{\omega E_{n}\left( S_{a}+E_{a}+A_{a}+R_{a} \right)\Delta PH(-\Delta P)}{N}], \\ &{E_{a}}^{'}(t)=q\lambda S_{a}-\sigma E_{a}-\frac{1}{\alpha}[\frac{\omega E_{a}\left( S_{n}+E_{n}+A_{n}+R_{n} \right)\Delta PH\left( \Delta P \right)}{N}+\frac{\omega E_{n}\left( S_{a}+E_{a}+A_{a}+R_{a} \right)\Delta PH\left( -\Delta P \right)}{N}], \\ &{I(t)}^{'}=\varrho\sigma E_{n}+\varrho\sigma E_{a}-\left( \delta_{I}\left( t \right)+\gamma_{I} \right)I, \\ &{A_{n}\left( t \right)}^{'}=\left( 1-\varrho\right)\sigma E_{n}-\gamma_{A}A_{n}+\frac{1}{\alpha}[\frac{\omega A_{a}\left( S_{n}+E_{n}+A_{n}+R_{n} \right)\Delta PH(\Delta P)}{N}+\frac{\omega A_{n}\left( S_{a}+E_{a}+A_{a}+R_{a} \right)\Delta PH(-\Delta P)}{N}], \\ &{A_{a}\left( t \right)}^{'}=\left( 1-\varrho\right)\sigma E_{a}-\gamma_{A}A_{a}-\frac{1}{\alpha}[\frac{\omega A_{a}\left( S_{n}+E_{n}+A_{n}+R_{n} \right)\Delta PH(\Delta P)}{N}+\frac{\omega A_{n}\left( S_{a}+E_{a}+A_{a}+R_{a} \right)\Delta PH(-\Delta P)}{N}], \\ &{H\left( t \right)}^{'}=\delta_{I}\left( t \right)I-\left( \alpha\left( t \right)+\gamma_{H} \right)H, \\ &{R_{I}\left( t \right)}^{'}=\gamma_{I}I+\gamma_{H}H, \\ &{R_{n}\left( t \right)}^{'}=\gamma_{A}A_{n}+\frac{1}{\alpha}[\frac{\omega R_{a}\left( S_{n}+E_{n}+A_{n}+R_{n} \right)\Delta PH\left( \Delta P \right)}{N}+\frac{\omega R_{n}\left( S_{a}+E_{a}+A_{a}+R_{a} \right)\Delta PH\left( -\Delta P \right)}{N}], \\ &{R_{a}\left( t \right)}^{'}=\gamma_{A}A_{a}-\frac{1}{\alpha}\left[ \frac{\omega R_{a}\left( S_{n}+E_{n}+A_{n}+R_{n} \right)\Delta PH\left( \Delta P \right)}{N}+\frac{\omega R_{n}\left( S_{a}+E_{a}+A_{a}+R_{a} \right)\Delta PH\left( -\Delta P \right)}{N} \right], \\ &{M\left( t \right)}^{'}=\eta\delta_{I}\left( t \right)I-\nu M. \end{aligned} \right.$ （S6）

where $H\left( \cdot\right)$ is the Heaviside function and $H\left( \Delta P \right)=1$when $\Delta P\geq0$ and $H\left( \Delta P \right)=0$when $\Delta P<0$.

Define$B=\left( S_{n}+E_{n}+A_{n}+R_{n} \right)/(S+E+A+R_{A})$, where $S=S_{n}+S_{a}, E=E_{n}+E_{a},A=A_{n}+A_{a},R_{A}=R_{n}+R_{a}$. Then by assuming that $\frac{S_{n}}{S}=\frac{E_{n}}{E}=\frac{A_{n}}{A}=\frac{R_{n}}{R_{A}}=B$, Model (S6) can be reduced to the following co-evolution model of the transmission dynamics and the behavioral change dynamics:

$\left\{ \begin{aligned} &S'=-\lambda\left[ B+q\left( 1-B \right) \right]S, \\ &E^{'}=\lambda\left[ B+q\left( 1-B \right) \right]S-\sigma E, \\ &I^{'}=\varrho\sigma E-\left( \delta_{I}\left( t \right)+\gamma_{I} \right)I, \\ &A^{'}=\left( 1-\varrho\right)\sigma E-\gamma_{A}A, \\ &H^{'}=\delta_{I}\left( t \right)I-\left( \alpha\left( t \right)+\gamma_{H} \right)H, \\ &R_{I}'=\gamma_{I}I+\gamma_{H}H, \\ &R_{A}'=\gamma_{A}A, \\ &M^{'}=\eta\delta_{I}\left( t \right)I-\nu M, \\ &B^{'}=\frac{\rho B\left( 1-B \right)\left( 1-mM \right)\left( S+E+A+R_{A} \right)}{N}, \end{aligned} \right.$(S7)

where $\rho=\frac{k\omega}{\alpha}, m=(m_{n}-m_{a})/k$ and $\lambda=\frac{\beta_{I}I+\left[ B+q\left( 1-B \right) \right]\beta_{A}A}{N}.1-mM$represents the balance between payoffs associated with two behaviors, and $1/m$ defines a threshold determining which behavior is more beneficial, and $\rho$ can be recognized as the speed of spontaneous behavioral changes with respect to transmission dynamics.

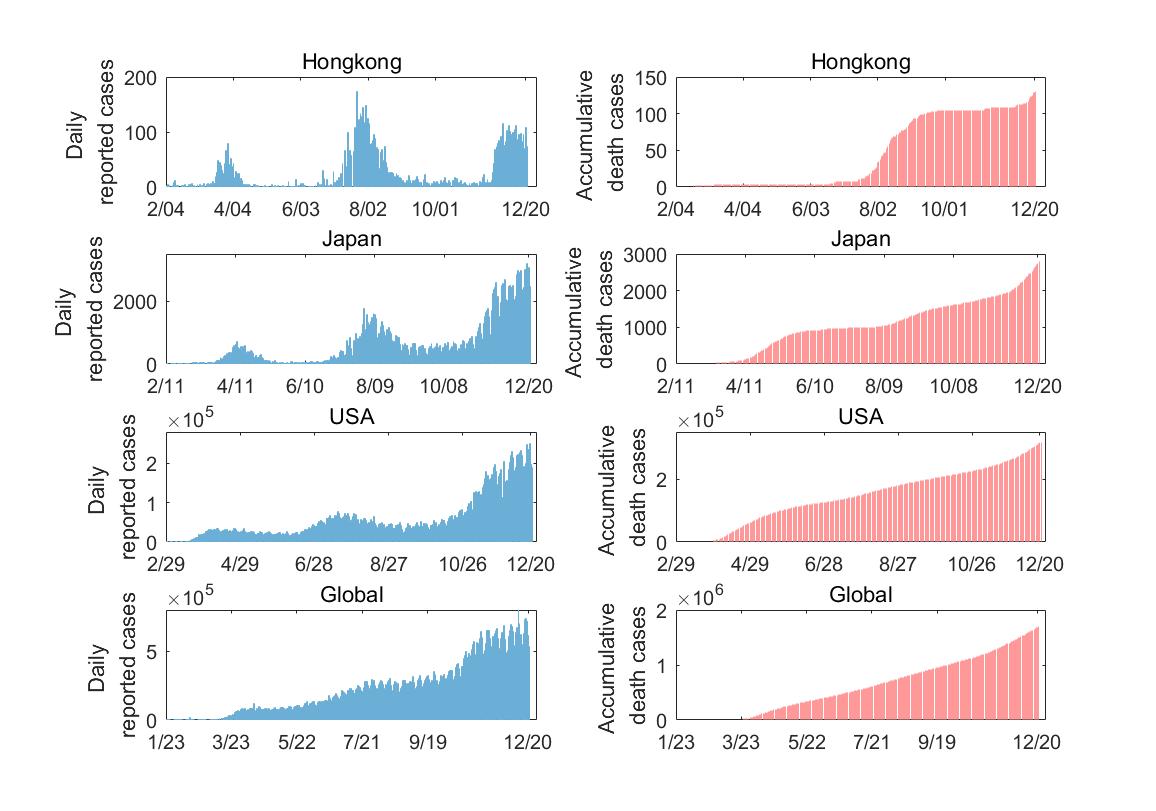

**SI Fig. 1** COVID-19 epidemic data, including the daily reported cases and accumulative death cases for Hongkong, Japan, USA and the world from the Johns Hopkins University Center for Systems Science and Engineering (JHU CCSE) and the data are available on the Github and the Humanitarian Data Exchange [28,29].

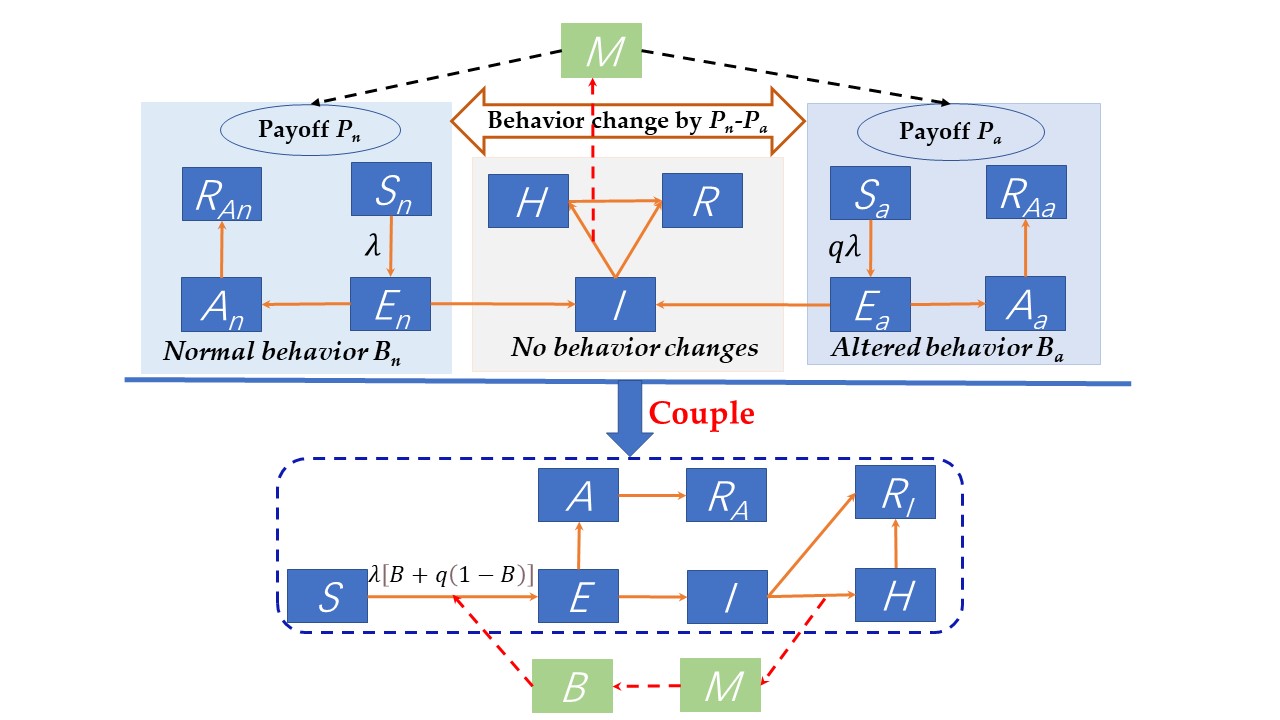

**SI Fig. 2** Schematic diagram for coupling the COVID-19 transmission dynamics and behavioral change dynamics.

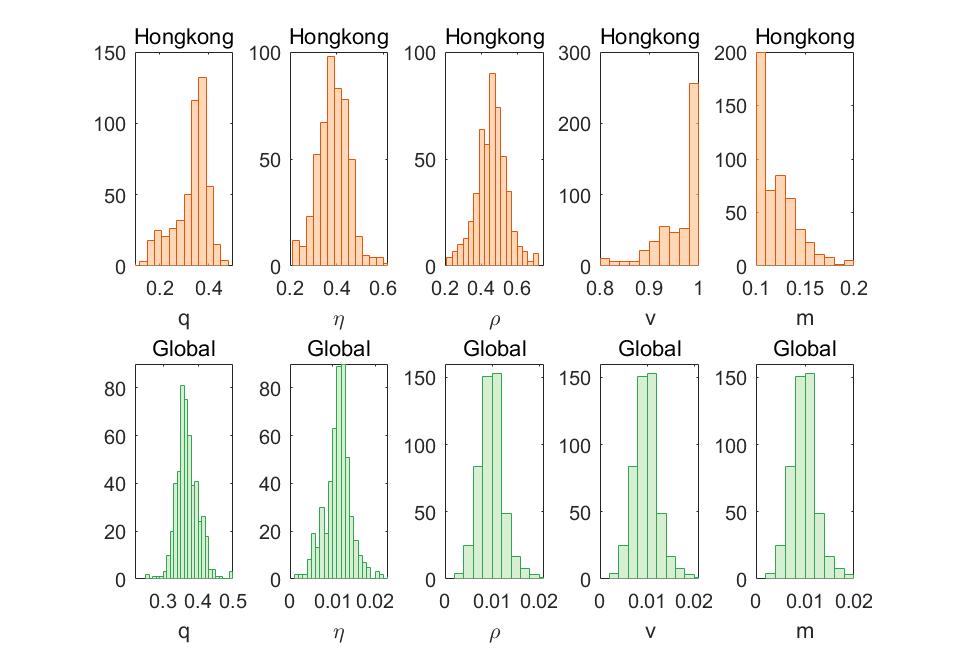

**SI Fig. 3** Empirical distributions of the estimated behavioral dynamic parameters by fitting Model (1) to the observed epidemic data from Hongkong and the world based on 500 bootstrap samples.

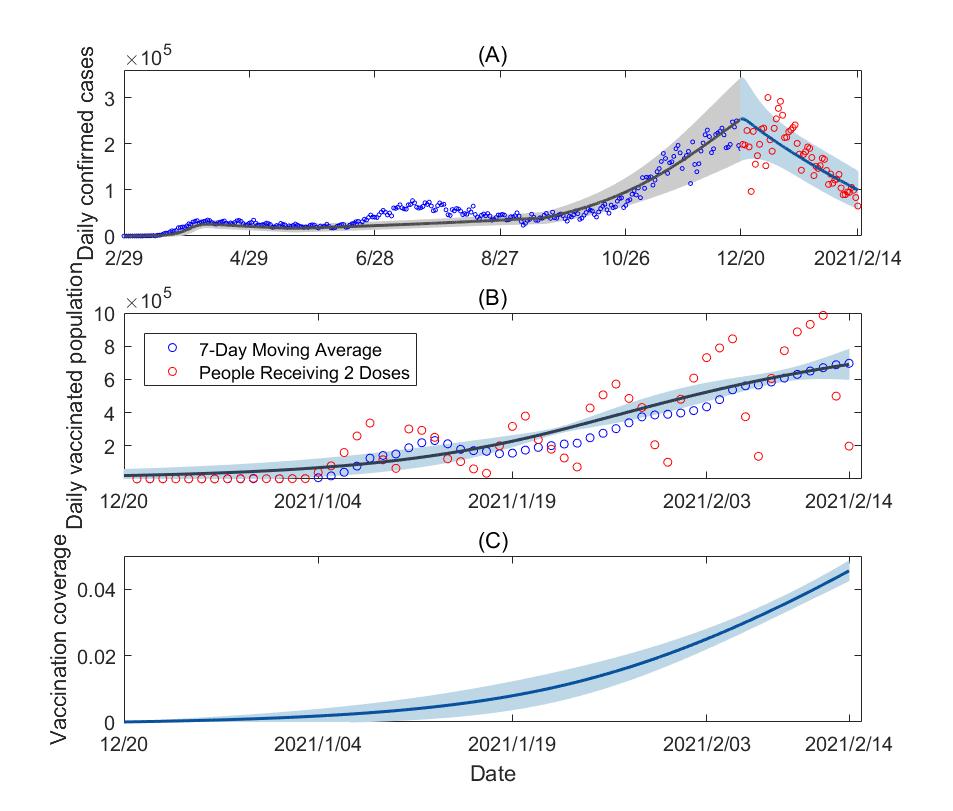

**SI Fig. 4**. (A)-(B) Model fitting results for the vaccination model (2) to the data between Dec 20, 2020 to Feb 14, 2021 in USA. (C) The estimated time-varying vaccination coverage in USA. The read cycles are the observed data, the blue curves are the estimated curves with the shadows as the corresponding 95% confidence interval from the bootstrap method.
